# Effects of latency and age structure on the dynamics and containment of COVID-19

**DOI:** 10.1101/2020.04.25.20079848

**Authors:** K.B. Blyuss, Y.N. Kyrychko

**Affiliations:** Department of Mathematics, University of Sussex, Falmer, Brighton, BN1 9QH, UK

## Abstract

In this paper we develop an SEIR-type model of COVID-19, with account for two particular aspects: non-exponential distribution of incubation and recovery periods, as well as age structure of the population. For the mean-field model, which does not distinguish between different age groups, we demonstrate that including a more realistic Gamma distribution of incubation and recovery periods may not have an effect on the total number of deaths and the overall size of an epidemic, but it has a major effect in terms of increasing the peak numbers of infected and critical care cases, as well as on changing the timescales of an epidemic, both in terms of time to reach the peak, and the overall duration of an outbreak. In order to obtain more accurate estimates of disease progression and investigate different strategies for introducing and lifting the lockdown, we have also considered an age-structured version of the model, which has allowed us to include more accurate data on age-specific rates of hospitalisation and COVID-19 related mortality. Applying this model to three comparable neighbouring regions in the UK has delivered some fascinating insights regarding the effect of quarantine in regions with different population structure. We have discovered that for a fixed quarantine duration, the timing of its start is very important in the sense that the second epidemic wave after lifting the quarantine can be significantly smaller or larger depending on the specific population structure. Also, the later the fixed-duration quarantine is introduced, the smaller is the resulting final number of deaths at the end of the outbreak. When the quarantine is introduced simultaneously for all regions, increasing quarantine duration postpones and slightly reduces the epidemic peak, though without noticeable differences in peak magnitude between different quarantine durations.

## 1 Introduction

As of 22nd April 2020, there are 133, 495 confirmed cases of coronavirus disease (COVID-19) and 18, 100 deaths reported in the UK, with more than 2.5M cases and almost 180, 000 deaths globally. The virus is reported to first appeared in Wuhan, China in 2019 [1], and has since silently and swiftly spread around the globe infecting populations in 212 countries and territories. When WHO declared it a global pandemic on the 11th of March 2020, there were 118, 000 cases in 100 countries. In order to contain and halt the spread of this deadly disease, countries around the world have taken extraordinary measures, introducing “lockdowns” and closing borders. The majority of cases of COVID-19 are spread via respiratory routes, especially among large gatherings of people in shopping malls, carnivals, celebrations etc., but a preliminary study has suggested that it can also spread via extra-respiratory routes, although the study was relatively small [2]. Clinically, COVID-19 is characterised by high temperature, cough and loss of smell and taste in some people, with cases varying from mild to very severe with life-threatening implications. The disease can also present itself with no symptoms at all, whereby someone infected with COVID-19 experiences no symptoms but is able to spread it to other people. It is thought that the number of asymptomatic carriers varies from 15 to 75 *–* 80%. While testing of the Diamond Princess Cruise ship has estimated that there were 17.9% of asymptomatic carriers [3], the population-wide testing of Vó Euganeo located 50 km west of Venice and closed off by authorities in February indicated that 50 *–* 75% of confirmed cases were symptomless [4]. This is one of the most challenging factors, which can have major implications in terms of managing and stopping the spread of the disease.

Another key epidemiological feature of COVID-19 is its relatively long incubation period, estimated to be about 5 days, where a person is exposed to the disease but the onset of symptoms appears some days later [5]. Since the exposed person can infect others while incubating the disease, this plays a major role in terms of epidemic control and management. The mortality rates from COVID-19 tend to be higher for older people, and children are thought to have either mild or no symptoms at all. CDC report of 6 April 2020 estimates that infection among people younger than 18 is 1.7%, 5.7% required hospitalisation compared to 10% for 18 *–* 64 age group [6], while hospitalisation rates in adults ages 80+ are estimated to be around 30 *–* 70% [13].

At the moment, in order to ease the strain on healthcare and save lives, majority of countries around the globe are practicing physical distancing/self-isolation measures, which are aimed at slowing the disease progressing and its intensity. Mathematical models have been widely used to analyse various scenarios of COVID-19 disease development, and to predict the best possible outcomes depending on the severity and length of introduced physical distancing measures [7, 8, 12, 19]. A number of these and other models [14, 15, 16] have used SIR- and SEIR-type epidemic models with an underlying assumption of exponential distributions of infection and recovery times.

In this paper we look into three specific aspects of COVID-19 dynamics and its containment. The first concerns an observation that the incubation period has a distinctly non-exponential distribution [5], and the same applies to infectious period [13]. We will include this feature in our model by means of a gamma distribution that much more accurately describes the behaviour of these major characteristics of disease dynamics. As we will show, this has a profound effect on disease dynamics in terms of its timescales (time to reach the peak and overall duration), as well as on the disease severity represented by the maximum number of infected individuals and the maximum number of critical care cases. The second aspect we will investigate concerns on observation that the infection, severity and mortality rates for COVID-19 are significantly different for different age groups [8, 12], thus it is essential to include the specific demographic structure of each particular region when model and assessing its potential needs for healthcare facilities, and, in particular, the number of critical care beds at different stages of epidemic progression. The third and final aspect we are interested in is the analysis of the effects of timing and duration of quarantine on containment of COVID-19 progression for regions with different demographic age structure.

The outline of the paper is as follows. In the next section we derive and analyse a mean-field model of disease dynamics with account for non-exponential distributions of incubation and infectious periods. In Section 3 we develop an age-structured version of this model that takes into account age-specific values of parameters and a demographic structure. Interactions between different age groups are modelled using three different types of age-specific mixing matrices for the UK population [9, 10, 20], including one obtained very recently after the lockdown was introduced in the UK. To illustrate the effects of different mixing patterns and quarantine, we will compare the results for three regions of the UK, focusing on the role of quarantine timing and duration.

## 2 Mean-field SEIR model

Before deriving a model for the dynamics of COVID-19, we note that one of the major assumptions behind SIR and SEIR-type models is an exponential distribution of incubation and recovery periods. Importantly, the actual distributions of these parameters as obtained from available epidemiological data rather obey a Gamma distribution, as illustrated in Fig. 1. To account for this fact in the model, we will represent Gamma distribution by means of models with multiple stages in the exposed and infected class, in a manner similar to Lloyd [17]. A very recent work Boldog et al. [18] considered an SEIR-type model of COVID-19 that explicitly includes two stages for exposed individuals and three stages for the infected class. However, the shape of distributions of incubation and recovery periods shown in Fig. 1 suggests that those numbers of stages may not be sufficient to properly represent the distributions of incubation and recovery periods; more specifically, with just two stages for exposed individuals, the distribution would be concave for smaller values of incubation period, while it should be convex according to the data from [5]. With these observations in mind, we model disease dynamics using a modification of the very recent model in Kissler et al. [19]: Susceptibles (*S*) get exposed to the disease, and after acquiring infection at rate *β* from infected individuals, they move to Exposed (*E*) class, stay there for an incubation period 1*/ν* before becoming infective (either symptomatically or asymptomatically), at which point a proportion *p*_*A*_ of them will move into Asymptomatic infected (*A*), a proportion *p*_*R*_ will move into Infected with mild symptoms (*I*_*R*_) not needing hospital and proceeding directly to recovery (*R*_*R*_), a proportion *p*_*H*_ will move into a class who will need hospital treatment (*I*_*H*_), and a proportion *p*_*C*_ will move into a class of those who will need critical care (*I*_*C*_). One additional assumption we make at this point is that the incubation period, which is the time from infection to the onset of symptoms, is the same as the latent period, which is the time from infection to becoming infectious. After recovery, characterised by an average recovery period 1*/γ*, Asymptomatic individuals move into the Recovered class (*R*_*A*_), and similarly, individuals in the *I*_*R*_ group move into the *R*_*R*_ class upon recovery. Those who are infected and require hospitalisation move into class *H*_*H*_ and then proceed to the Recovered class *R*_*H*_ after an average period of 1*/δ*_*H*_. The infected class who will require critical care *I*_*C*_ move into the hospitalised class *H*_*C*_, from which they proceed to a critical care class *C*_*C*_ at rate *δ*_*C*_, and subsequently they either move into the recovered class *R*_*C*_ at rate *ξ*_*C*_, or die and move to a compartment *D* at rate *µ*. In terms of including the above-mentioned distributions of incubation and recovery periods in the model, we will assume that with the same mean incubation period, individuals in the Exposed class go through *K*_1_ sequential stages of equal duration, and in the Asymptomatic and Infected classes they go through *K*_2_ stages of the same duration. With these assumptions, the model equations now have the form

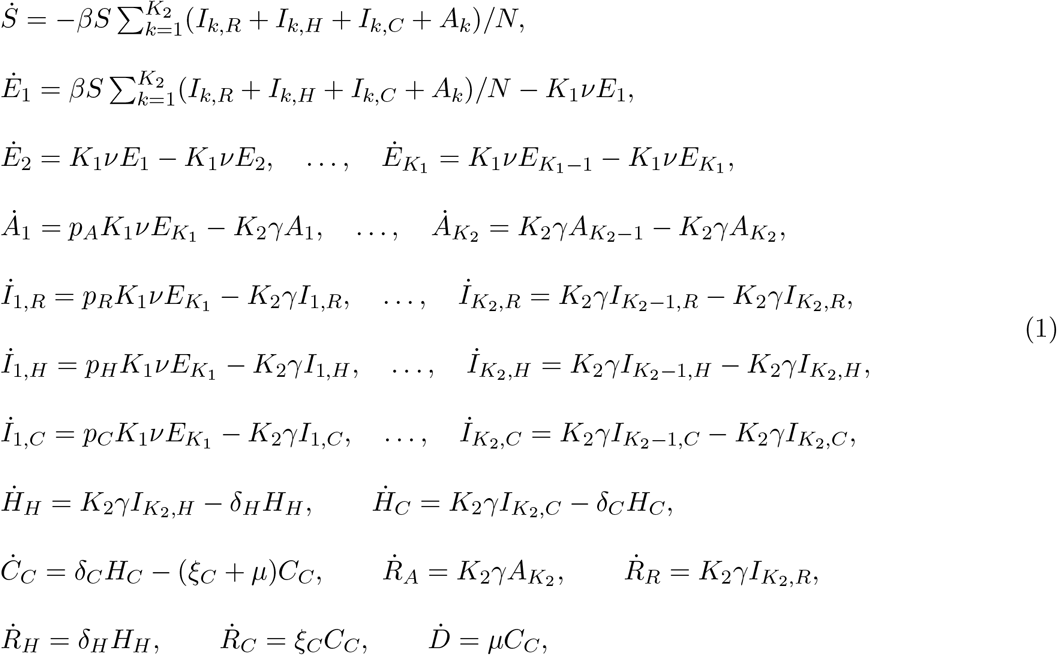

where dot denotes the derivative with respect to time, and the parameters are

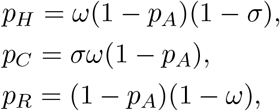

where *ω* is the rate of hospitalisation and *σ* is the rate of critical care admission [12, 13].

**Figure 1:**
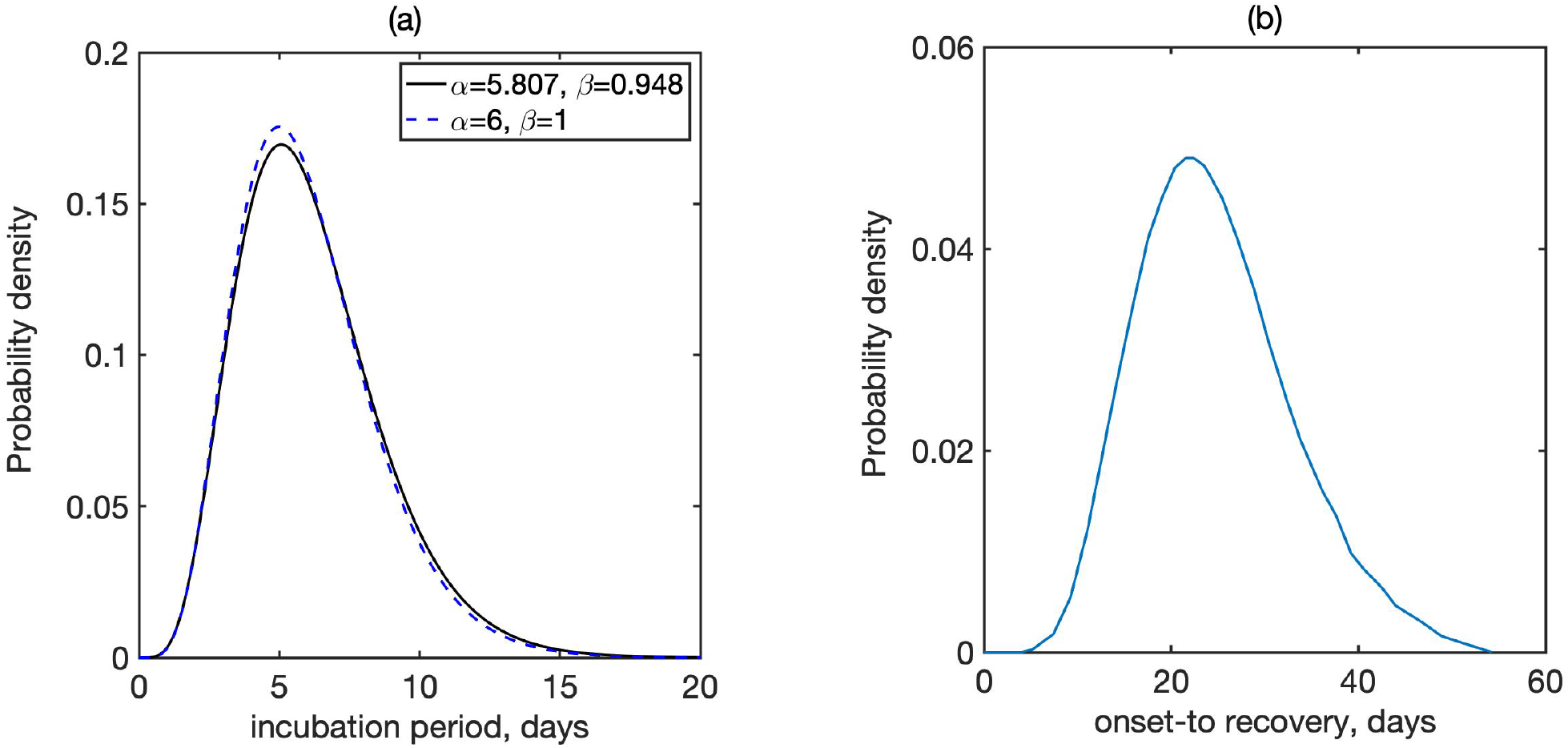
Distribution of (a) incubation period, where *α* is the shape and *β* is the scale of Gamma distribution, and (b) recovery periods [13, 5]. In (a), solid line shows the bets fit to data from Lauer et al. [5], the dashed line is the closest fit to Gamma distribution with integer parameters.

Figure 3 shows a comparison of time dynamics of the epidemics with baseline values of parameters from Table 1 and different numbers of stages of incubation and recovery. This figure shows that the overall dynamics changes drastically when the numbers of stages are varied, and the situation for *K*_1_ = *K*_2_ = 1, which corresponds to the standard SEIR model, is significantly different from the situation with *K*_1_ = *K*_2_ = 6, which provides a much more realistic representation of incubation and recovery periods illustrated in Fig. 1. We observe that for the same mean incubation and recovery periods, increasing the number of stages in the incubation period results in slightly bringing forward the peak of the epidemic and its overall completion, while increasing the number of stages in the recovery period significantly increases the maximum total numbers of infected and the number of critical cases.

**Table 1:** Parameter definitions and their baseline values from [19], except for *γ* from [5] and *β* adjusted to have *R*_0_ = 2.5.

| Parameter | Value | Meaning |
| --- | --- | --- |
| $\beta$ | 0.15 | disease transmission/infection rate |
| $K$ | 1 – 6 | number of exposed stages |
| $1/\nu$ | 5 days | incubation time |
| $1/\gamma$ | 16.6 days | infectious period/time to hospitalisation (both non-critical and critical care) |
| $p_A$ | 0.5 (assumed) | proportion of asymptomatic infections |
| $p_R$ | 0.456 (varies) | proportion of symptomatic infections without the hospital/critical care requirements |
| $p_H$ | 0.0308 (varies) | proportion of symptomatic infections requiring hospitalisation, but not critical care |
| $p_C$ | 0.0132 (varies) | proportion of symptomatic infections requiring critical care |
| $1/\delta_H$ | 8 days | recovery rate for non-critical hospital cases |
| $1/\delta_C$ | 8 days | average duration from hospital admission to requiring critical care |
| $1/\xi_C$ | 10 days | recovery rate for admissions requiring critical care |
| $\mu$ | 0.1 (varies) | death rate for admissions requiring critical care |

**Figure 2:**
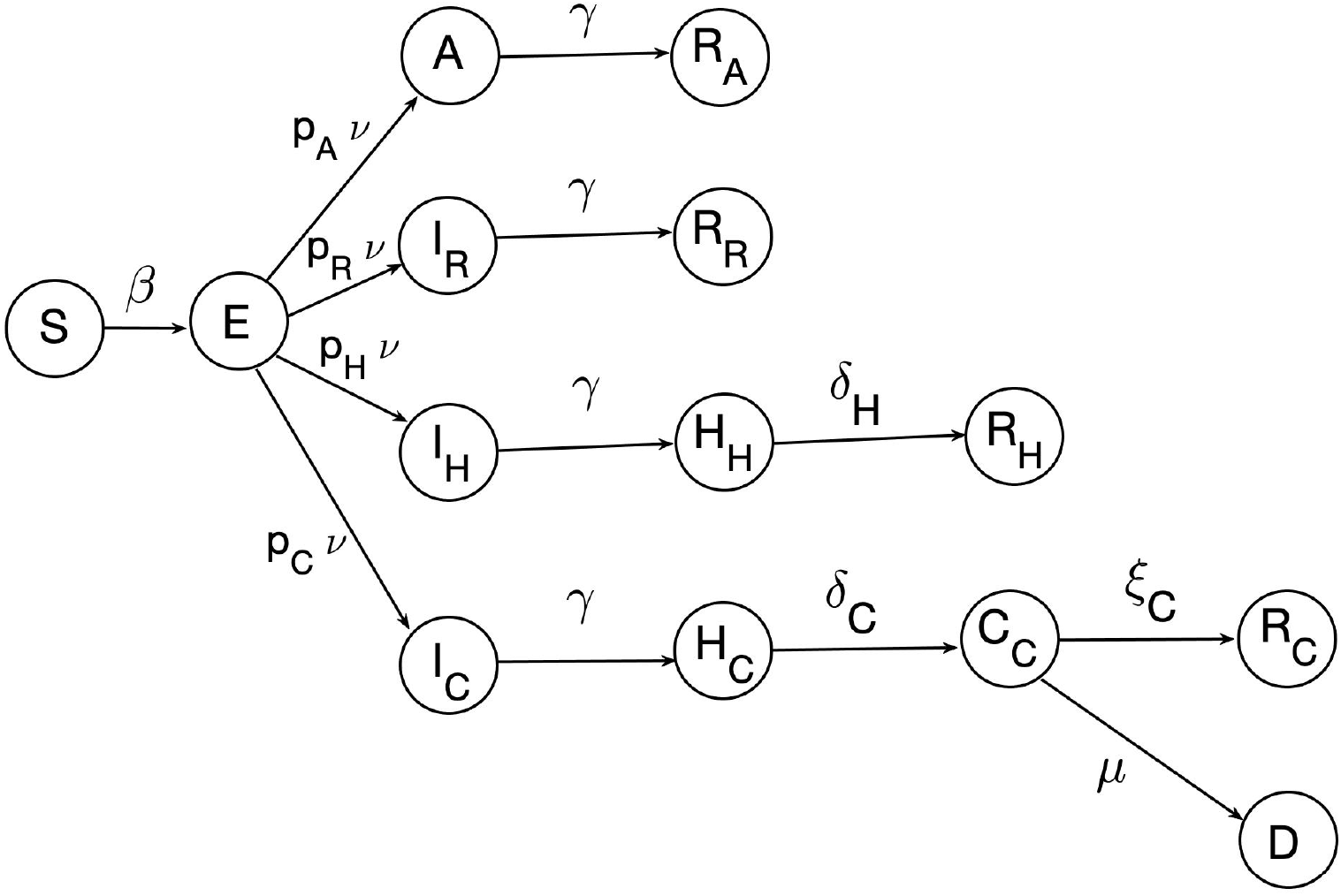
Schematic diagram of the disease transmission model.

**Figure 3:**
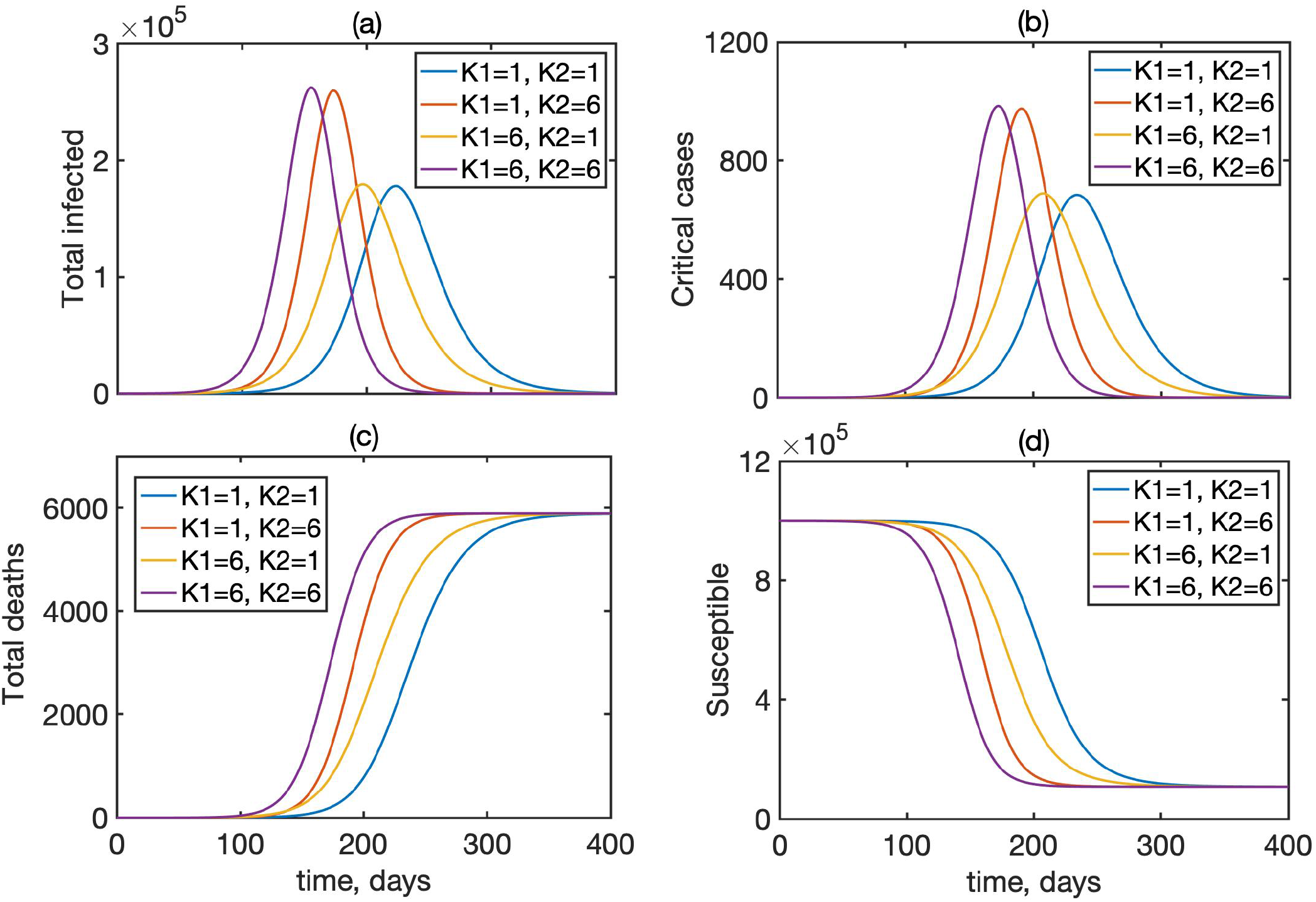
Dynamics of the total infected (*A* + *I*_*H*_ + *I*_*R*_ + *I*_*C*_), critical cases *C*, total deaths *D*, and the susceptibles *S* in model (1) for different numbers of incubation and recovery stages, with *N* = 1, 000, 000, and 5 individuals initially exposed to infection.

## 3 Age-structured model with latency and gamma distribution of incubation and recovery periods

An equivalent model that accounts for age distribution of parameters has the form

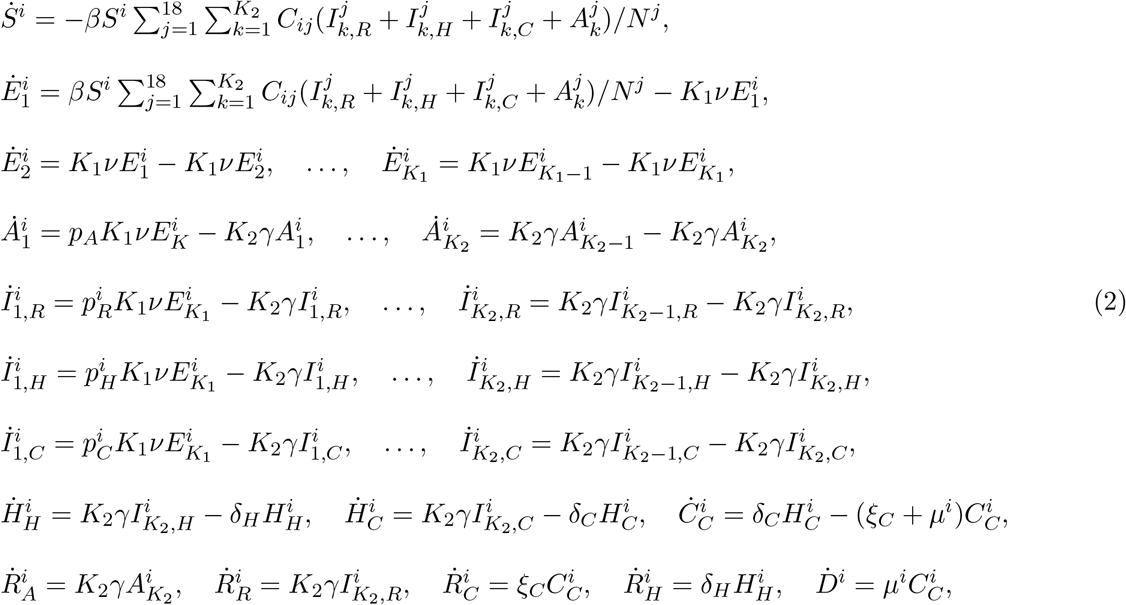

where *i* = 1‥18 are different age groups, *K*_1_ is the number of incubation stages and *K*_2_ is the number of recovery stages, *C*_*ij*_ is the mixing matrix. For simulations, it was assumed that a small proportion in each age group was initially exposed. Age-specific values of parameters *ω, σ* and *µ* used for simulations are given in Table 3 and are based on Ferguson et al. [12].

**Table 2:** Variables of the model.

| Variable | Meaning |
| --- | --- |
| $S$ | Susceptibles |
| $N$ | Total population size |
| $E_i, i = 1, \dots, K_1$ | Exposed individuals in different stages after the exposure to the disease |
| $A_i, i = 1, \dots, K_2$ | Asymptomatic infected individuals in different stages |
| $I_{R,i}, i = 1, \dots, K_2$ | Infected individuals in different stages, who will not require hospitalisation |
| $I_{H,i}, i = 1, \dots, K_2$ | Infected individuals in different stages, who will require hospitalisation |
| $I_{C,i}, i = 1, \dots, K_2$ | Infected individuals in different stages, who will require hospitalisation followed by critical care |
| $H_H$ | Hospitalised individuals not requiring critical care |
| $H_C$ | Hospitalised individuals requiring critical care |
| $C_C$ | Individuals in critical care |
| $R_A$ | Recovered asymptomatic individuals |
| $R_R$ | Recovered individuals without hospitalisation |
| $R_H$ | Recovered individuals after hospital (without critical care) admission |
| $R_C$ | Recovered individuals after critical care |
| $D$ | Those who died in critical care |

**Table 3:** Age-specific parameter values from [12].

| Age-group (years) | % symptomatic cases requiring hospitalisation, $\omega$ | % hospitalised cases requiring critical care, $\sigma$ | infection fatality ratio, $\mu$ |
| --- | --- | --- | --- |
| 0 to 9 | 0.1% | 5% | 0.002% |
| 10 to 19 | 0.3% | 5% | 0.006% |
| 20 to 29 | 1.2% | 5% | 0.03% |
| 30 to 39 | 3.2% | 5% | 0.08% |
| 40 to 49 | 4.9% | 6.3% | 0.15% |
| 50 to 59 | 10.2% | 12.2% | 0.6% |
| 60 to 69 | 16.6% | 27.4% | 2.2% |
| 70 to 79 | 24.3% | 43.2% | 5.1% |
| 80+ | 27.3% | 70.9% | 9.3% |

### 3.1 Mixing matrices

To model the effects of different levels of interaction between individuals in the population, we will consider three different mixing matrices. The first one, known as POLYMOD, comes from a major UK study in 2008 [9] and has been subsequently used to model the infectious disease dynamics in a variety of contexts. The second mixing matrix was produced as a result of UK’s nationwide social experiment on online tracking of an influenza epidemic for a BBC programme “Contagion!” in 2018 [10, 11]. The BBC mixing matrices (all and physical only) were padded with POLYMOD data for the missing block describing interactions in the lowest age groups, which were scaled to ensure that the leading eigenvalue of the BBC matrices did not change. The resulting mixing matrices are largely similar to the POLYMOD matrices, with the only main difference being a change in the structure of interactions between teenagers and other age groups. This change was largely attributed to a significant level of using smartphones and other gadgets by teenagers, which resulted in the reduction of direct social interactions for this age group. The third matrix we will use comes from the so-called CoMix survey [20], an online survey conducted in March 2020 by Ipsos that looked into levels of social mixing in the UK following the introduction of a lockdown.

Since the POLYMOD and BBC matrices describe regular interactions between different age groups in the absence of any intervention, they were scaled with their respective leading eigenvalue to maintain the same basic reproduction number for the same transmission rate *β*. When analysing the effects of quarantine, we will use BBC-all mixing matrix as a baseline, with CoMix-all describing contacts during the quarantine, and for the same transmission rate, the CoMix matrix is rescaled by a leading eigenvalue of the BBC-all matrix to reflect a reduction in the number of contacts between these two matrices. As discussed in Jarvis et al. [20], for a baseline value of the basic reproduction number of *R*_0_ = 2.6, under CoMix mixing, this value was reduced to 0.62 for the case of all contacts, and to 0.37 for physical contacts only.

### 3.2 Age-specific regional information

To explore the dynamics of COVID-19 epidemics and its control using quarantine in regions with difference social structure, we will consider three neighbouring coastal UK regions, namely, West Sussex, Brighton and Hove, and East Sussex. These regions have respective total populations of 858,700, 290,500 and 554,800, median ages 41.1, 35.6 and 42.7 [21], and their age distributions are illustrated in Figure 5.

**Figure 4:**
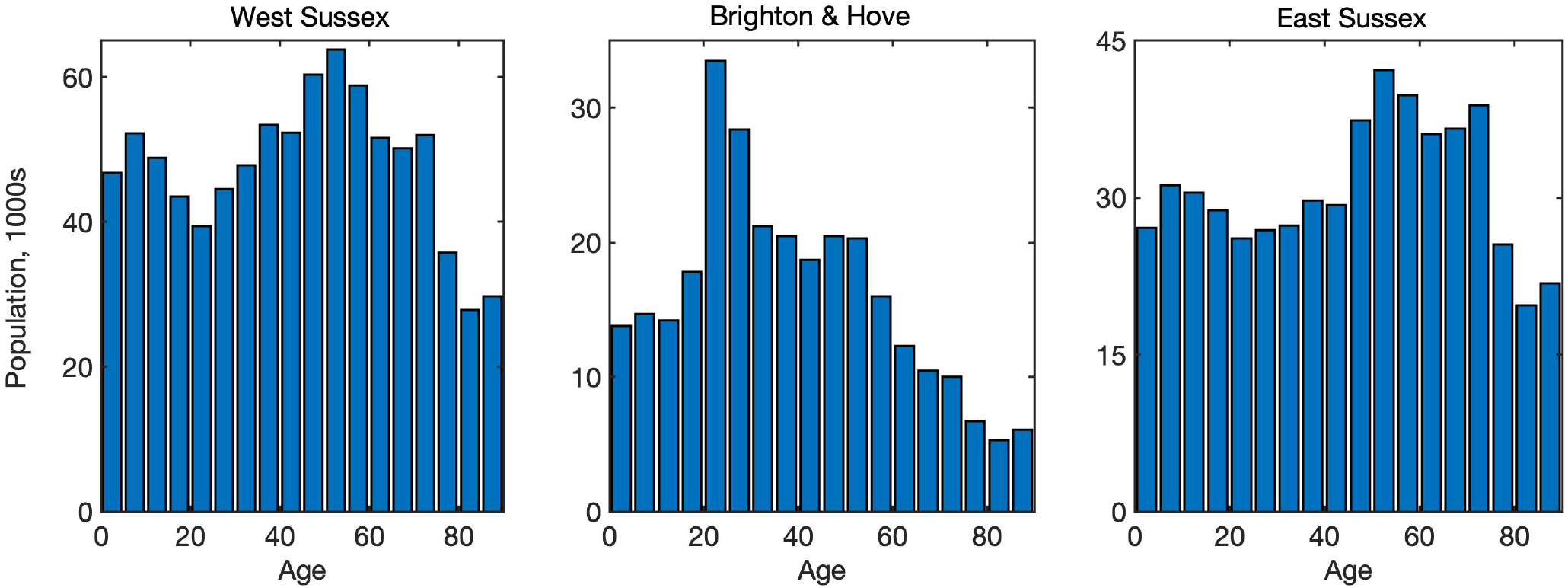
Age distribution in West Sussex, Brighton and Hove, and East Sussex [21].

**Figure 5:**
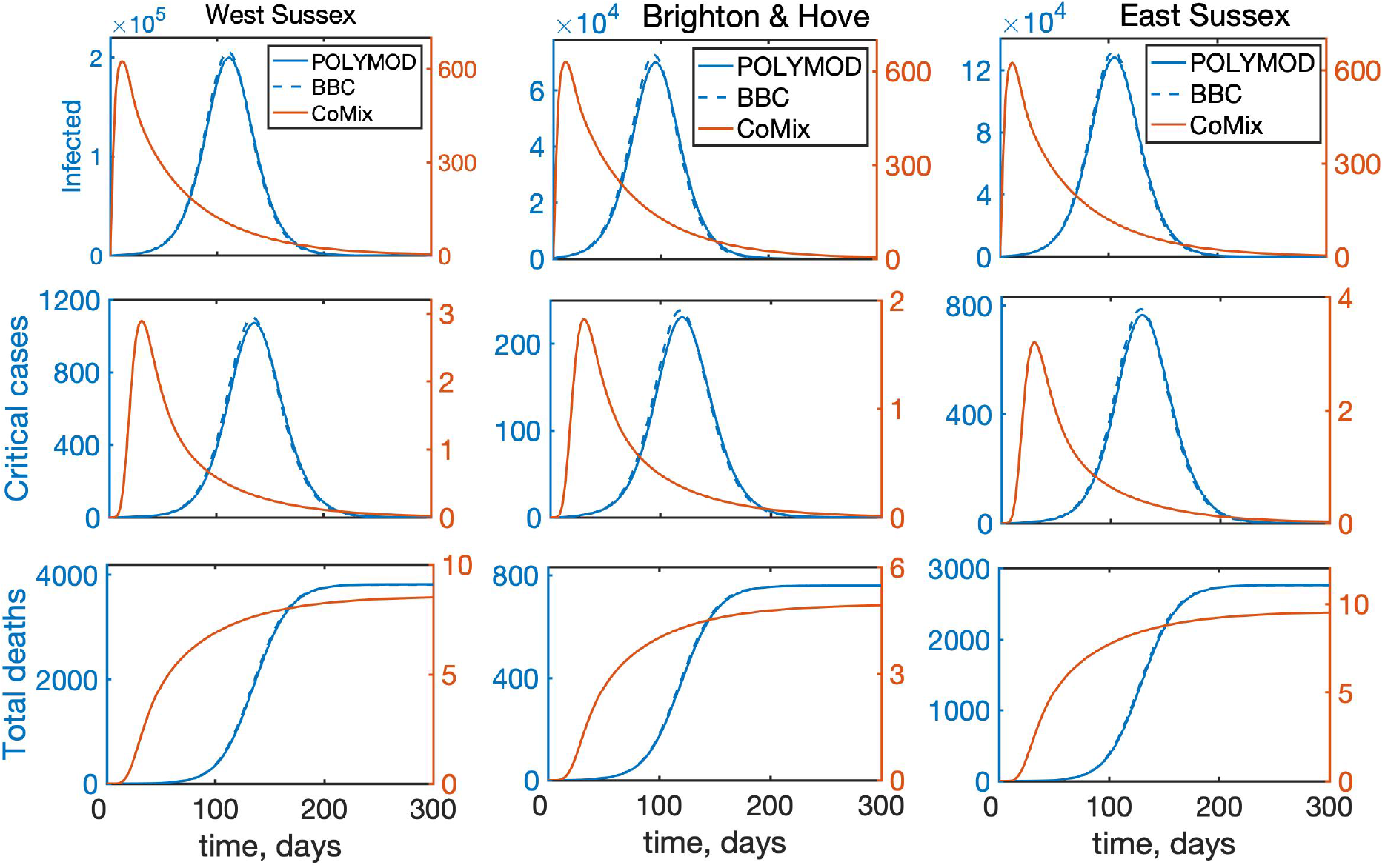
Temporal dynamics of the model (2) with all interactions between individuals for West Sussex, Brighton and Hove, and East Sussex. “Infected” denotes the total number of infected individuals, including asymptomatic, symptomatic, and all who require hospitalisation.

## 4 Results

For all numerical simulations in this section, we have used the values of parameters as given in Tables 1 and 3, with *K*_1_ = *K*_2_ = 6, and the resulting model (2) was solved numerically using a Runge-Kutta-Fehlberg method. As an initial condition we took 100 people incubating the disease, distributed among age classes with proportions being the proportions in each age class, as determined by the actual age distribution for each specific region.

### 4.1 Effects of mixing matrices and age structure

To investigate the role of interactions between different age groups in the dynamics of COVID-19, we have solved the model (2) numerically using each of the three mixing matrices, assuming that they characterise interactions between individuals from the very start of an epidemic until its end. Figure 5 shows the results of such simulations for the cases, where **all types** of contacts were included, and the corresponding age distribution of deaths at the end of the epidemic is illustrated in Fig. 6. We observe that for the more recently obtained BBC-all mixing matrix, the numbers of infected and critical cases are very slightly greater than for the POLYMOD mixing, though the differences are negligibly small. In contrast, for the CoMix mixing, all these numbers are two orders of magnitude smaller, supporting the underlying idea that the CoMix mixing matrix describes a substantially reduced level of interactions between individuals during a quarantine. When one looks only at physical interactions between individuals, as shown in Fig. 7, the differences between POLYMOD-all and BBC-all mixing become more pronounced, with the numbers of infected and critical cases being higher for the BBC mixing compared to the POLYMOD mixing, though for the total deaths the differences are still very small. The age structure of deaths at the end of an epidemic is qualitatively the same for both all and physical contacts: the numbers of deaths are very small up until around 60 years of age, and after that they are growing, with the highest numbers of deaths observed in the eldest people, as consistent with current data. The underlying reason for this is that, as indicated in Table 3, the rates of hospitalisation, critical care, and fatality all increase with age and are highest for the age groups of 60+. We have explored how the dynamics would change if the initial conditions were to be modified, and the conclusion is that all results would remain effectively the same, with minor changes in terms of shifts in time and in magnitude.

**Figure 6:**
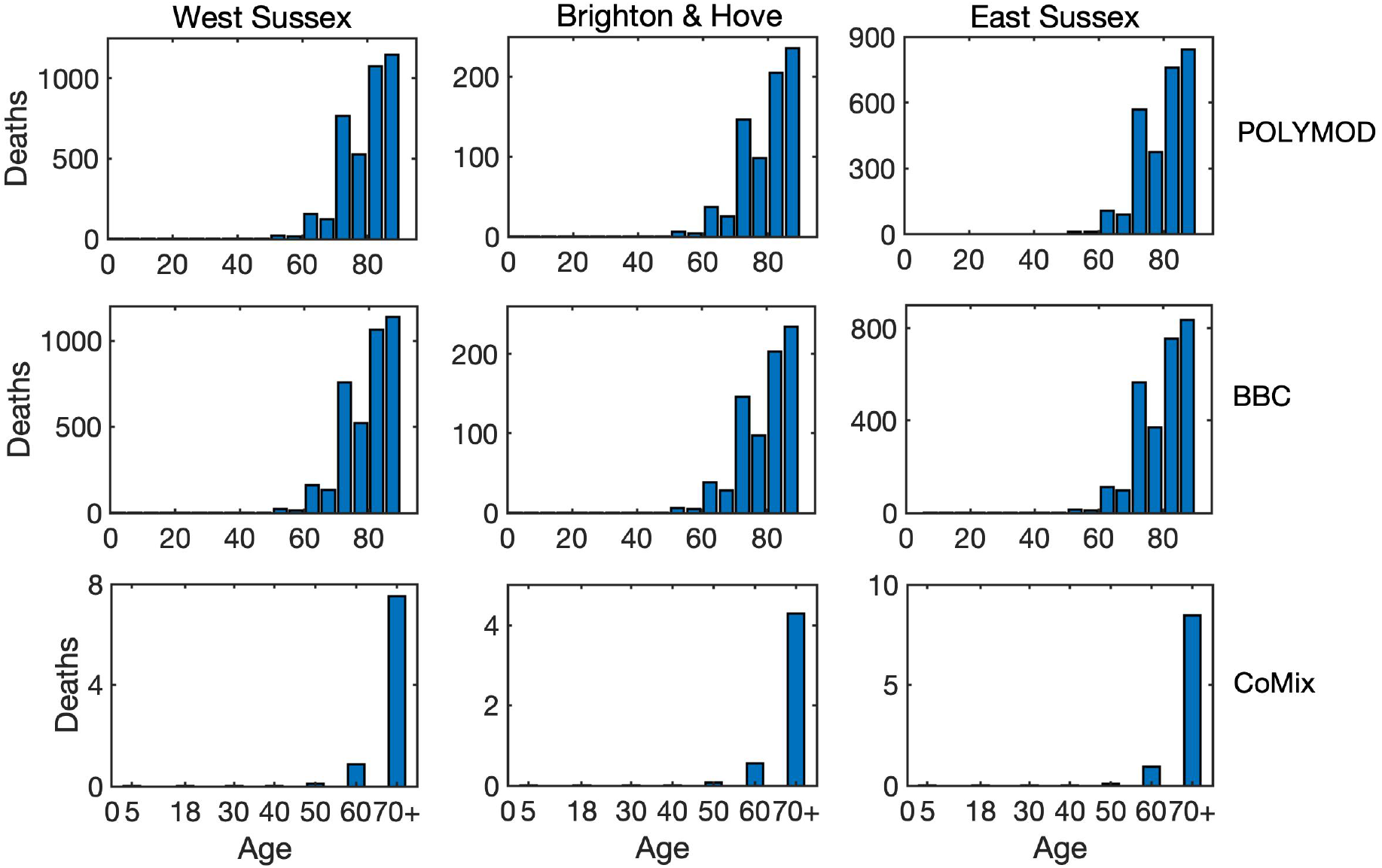
Age distribution of death cases at the end of an epidemic with all interactions between individuals for West Sussex, Brighton and Hove, and East Sussex.

**Figure 7:**
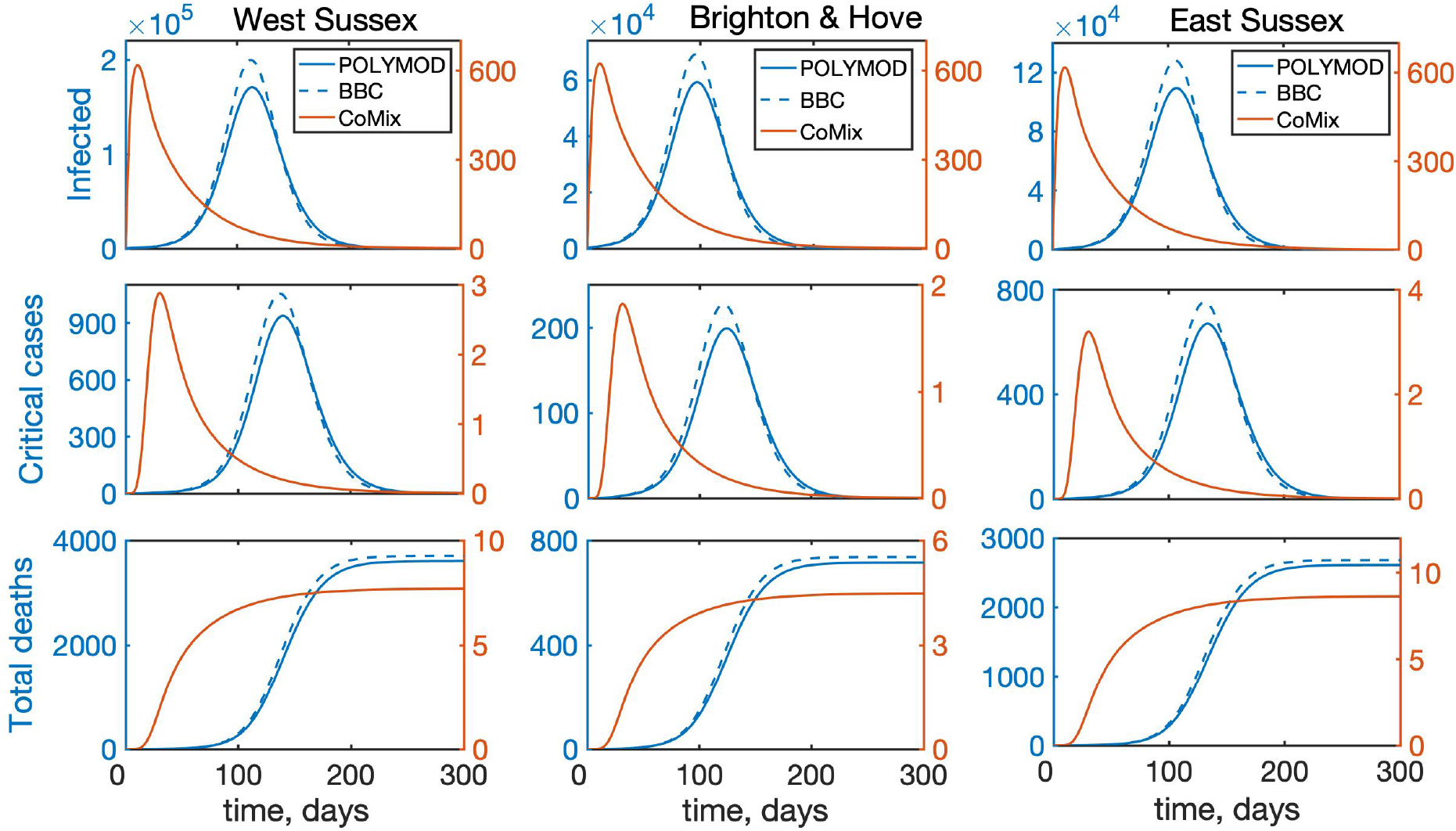
Temporal dynamics of the model (2) with account only for direct physical interactions between individuals for West Sussex, Brighton and Hove, and East Sussex. “Infected” denotes the total number of infected individuals, including asymptomatic, symptomatic, and all who require hospitalisation.

### 4.2 Effects of timing and duration of quarantine

As a next step, we have looked into the effect of introducing quarantine on disease dynamics. Simulations shown in the previous section describe what would happen if the quarantine were introduced from the very beginning of the epidemic and stayed in force for the entire duration of the outbreak. In reality, however, quarantine is normally introduced some time after the start of an outbreak and lifted after a certain period of time, with the conflicting requirements of trying to minimise the number of critical cases and deaths, while also trying to minimise the duration of the quarantine. To model this scenario, we have chosen BBC-all as the underlying matrix describing interactions in the absence of quarantine, and CoMix describing interactions during quarantine. Figure 9 shows how the disease dynamics changes depending on when the quarantine is introduced relative to the start of an epidemic outbreak, assuming that in all cases the quarantine remains in force for 8 weeks, after which point it is lifted. Although in each case there will be a second epidemic peak after the quarantine is lifted, interestingly and quite counter-intuitively, the **later** is the quarantine introduced, **the smaller** will be the maximum of the two peaks, and this reduction becomes quite substantial for quarantines introduced at a later stage. One possible explanation of this is that if the quarantine is introduced very early on into the outbreak, the number of people who actually have the disease is still relatively small, and as the epidemic goes through its course, if the quarantine is lifted closer to the peak of the epidemic, there will be a large number of infected people in the population, and lifting the quarantine will release a pool of additional susceptibles who can get a disease. In contrast, if the quarantine is introduced later, so that it is lifted after the “natural” peak of the epidemic, the numbers of infected people will already be significantly reduced, so the second peak will be much smaller. Another interesting observation is that for the same quarantine duration, the timing may also be important in determining which of the two epidemic peaks will be larger, and this is different for different regions. The reason for the difference between regions is that since the main driving force behind any disease transmission is interactions between individuals, and even for the same mixing matrix these are very strongly determined by the underlying population structure. Figure 9 indicates that for a fixed-duration quarantine introduced relatively early, i.e. 6 or 8 weeks after the start of an epidemic, in all three regions we have considered the first epidemic peak will be much smaller than the second peak. For a quarantine that is introduced after 10 weeks, the same conclusion still holds for West Sussex and East Sussex, which are both characterised by a significantly older population. In contrast, in Brighton and Hove the second peak is significantly smaller than the first, and this can be attributed to the fact that younger people have a chance to go through the cycle of disease earlier on in the epidemic, and since they are the biggest contributors to disease transmission due to a higher number of their contacts, when the quarantine is lifted, there will be fewer of them capable of transmitting the disease to the rest of the population. At the same time, a reduction in the total number of deaths by the end of en epidemic is much more strongly reduced for quarantines that are introduced later: it is 7.1-19.5% for an 8-week quarantine introduced after 10 weeks, compared to only 1-3% for a quarantine introduced after 6 weeks). A major implication of this result is that the timing of optimal introduction of a fixed-duration quarantine should be adjusted for each individual region to make it most effective in terms of reducing the number of infections and critical care cases, and to avoid exacerbating the outbreak after the quarantine is lifted. Alternatively, quarantine duration should be sufficiently large, so that it would substantially exceed the time required to reach a natural peak of an epidemic, so that a subsequent lifting of the quarantine would not result in a major resurgence of infections and critical care cases.

**Figure 8:**
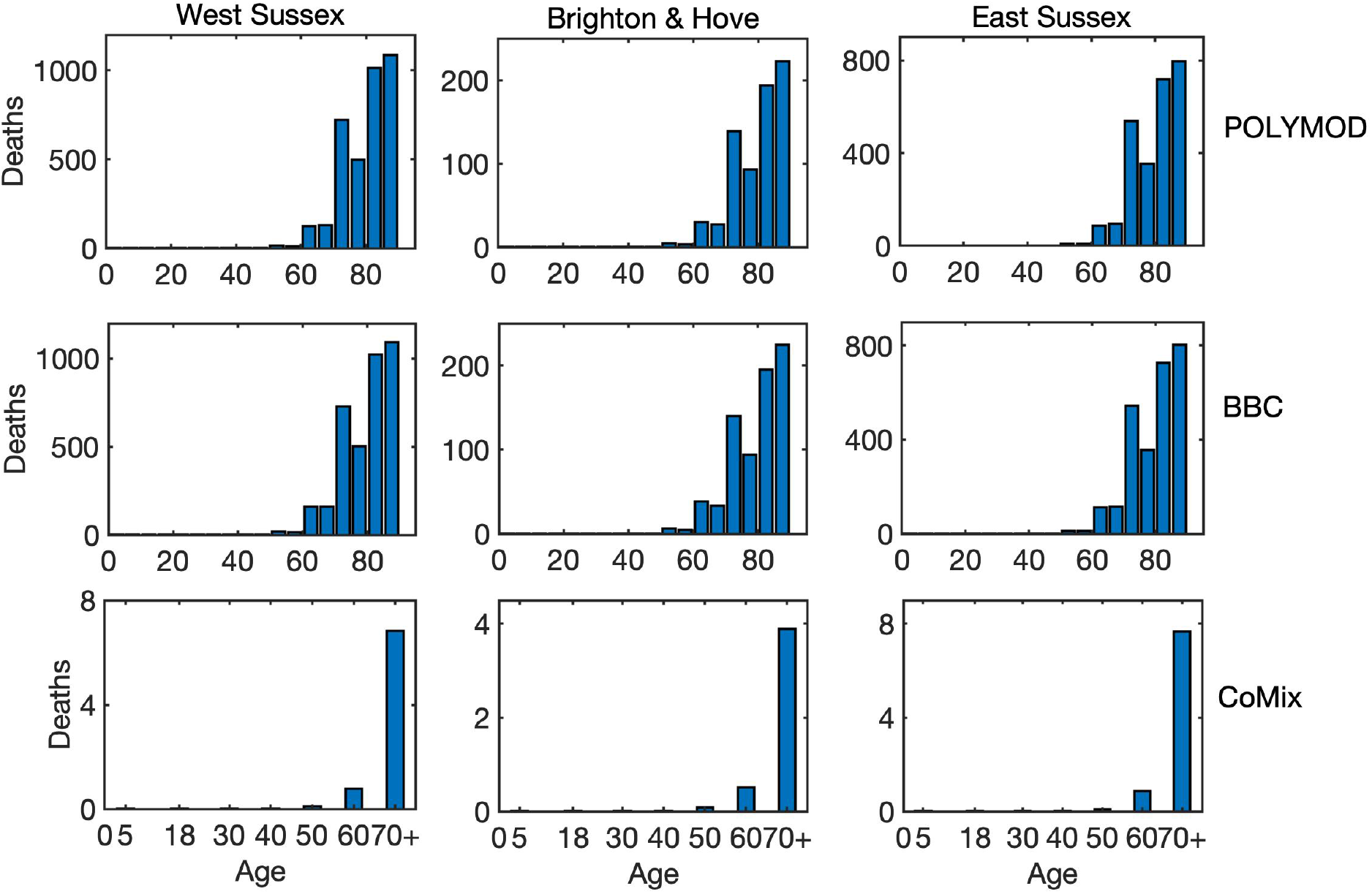
Age distribution of death cases at the end of an epidemic with only physical interactions between individuals for West Sussex, Brighton and Hove, and East Sussex.

**Figure 9:**
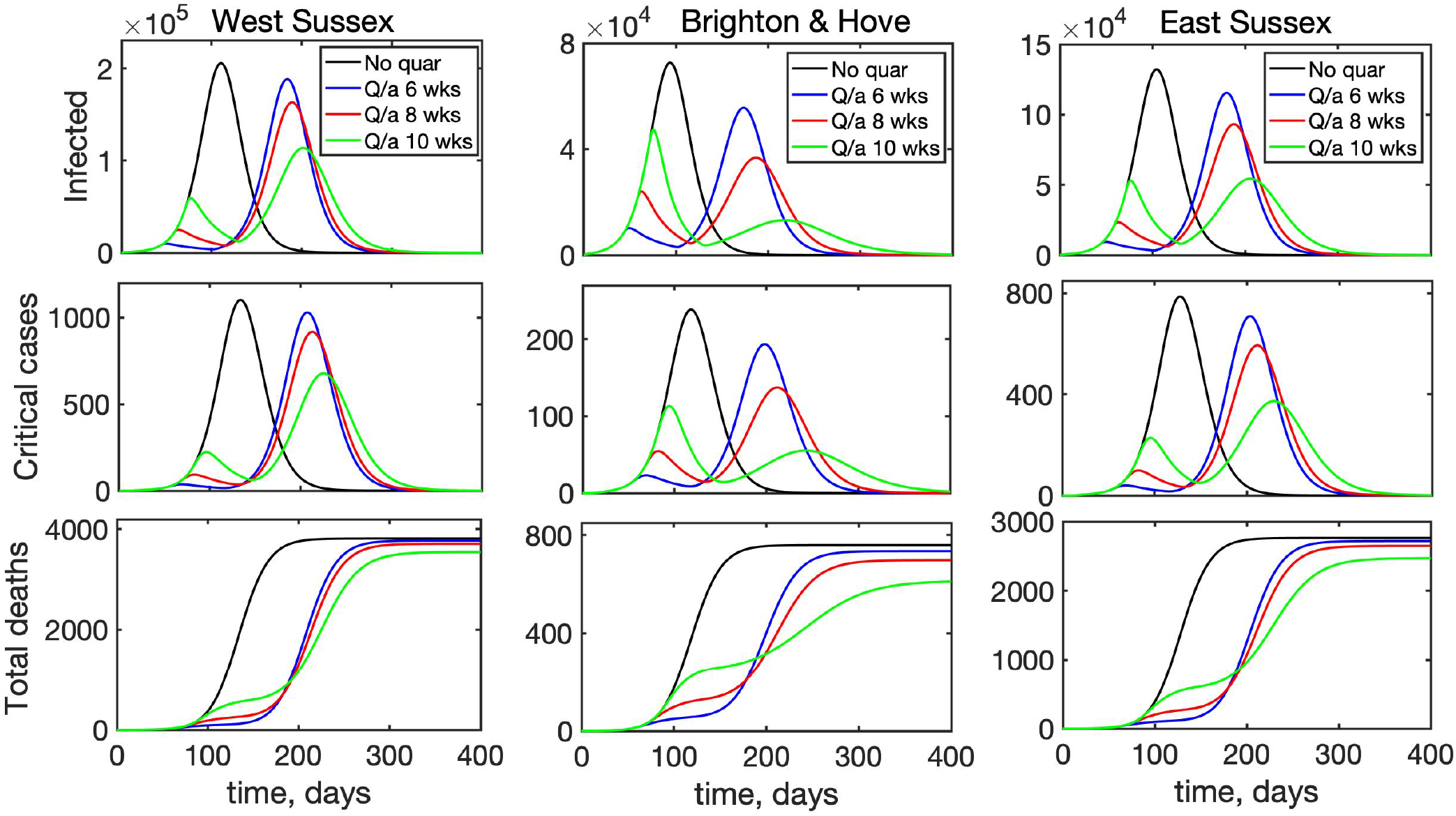
Effects of the time of introduction of quarantine on disease dynamics with BBC-all mixing matrix as baseline (black), and quarantine introduced for 8 weeks starting after 6 weeks (blue), 8 weeks (red), or 10 weeks (green).

Another question of major research and practical importance is when is most appropriate time to lift quarantine restrictions and to allow people to resume their work and social contacts. To investigate this, we have considered the following scenario: an epidemic starts in a population, whose interactions are described by the BBC-all mixing matrix, then after 8 weeks a quarantine is introduced, which is modelled by the CoMix-all matrix, and the quarantine stays in force for 8, 12, or 16 weeks. At this point the quarantine is lifted, and the population mixing returns to its original state described by the BBC-all mixing matrix. Figure 10 illustrates the resulting time dynamics of the epidemic, and it shows that in all cases when the quarantine is introduced, the curve is “flattened”, with the peak in the number of infected and deaths being smaller for longer quarantine duration. Since in all three simulated scenarios the quarantine is introduced relatively soon into the outbreak, the second epidemic peak is much higher than the first regardless of how long the quarantine lasts. Also, quite surprisingly, the reduction in the magnitude of the peak in the numbers of infected and critical care cases between an 8-week quarantine and a 16-week quarantine is quite small (5.4-7.4% for infected, and 5-8% for critical care cases), raising the question about practical justification of the longer quarantine. Another observation is that since the quarantine is introduced relatively early on, the difference between different regions becomes less pronounced, though similarly to the above analysis of the timing of the quarantine, we notice that a longer quarantine in Brighton and Hove makes the difference between two epidemic peaks much smaller, while both in West Sussex and East Sussex, the second peak remains significantly larger than the first for any quarantine duration. Interestingly, there are also almost no differences between the total numbers of deaths for different quarantine durations, though an overall reduction compared to a situation without quarantine is notably different for different regions: it is only 2.8-3.7% for West Sussex and 4.2-5.5% East Sussex, compared to 8-11% for Brighton and Hove.

**Figure 10:**
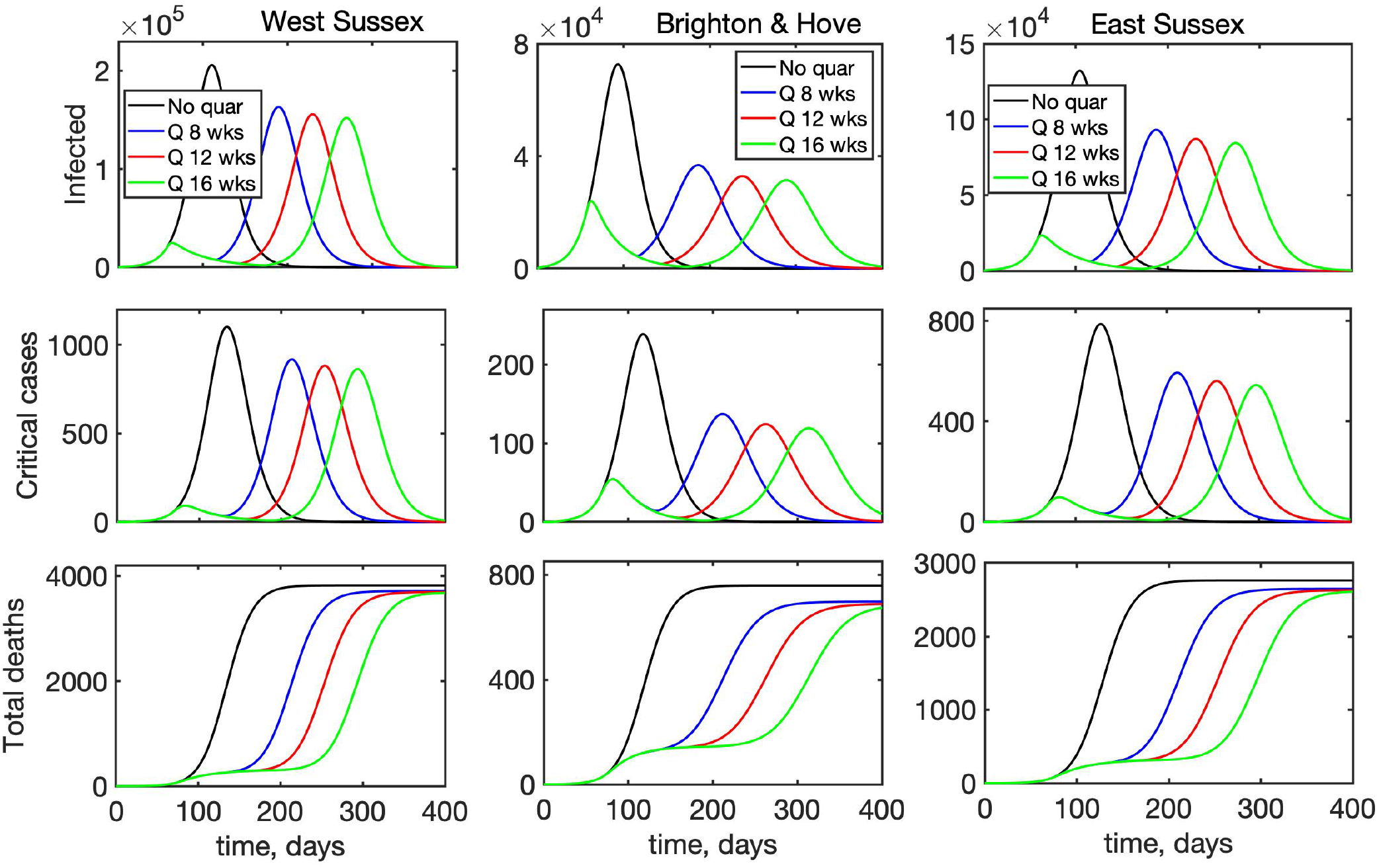
Effects of the duration of quarantine on disease dynamics with BBC-all mixing matrix as baseline (black), and quarantine introduced after 8 weeks for 8 weeks (blue), 12 weeks (red), or 16 weeks (green).

## 5 Discussion

In this paper we have considered the effects of non-exponential distributions of incubation and recovery periods which are suggested by available data, as well as age structure, on dynamics of COVID-19 and its possible containment using quarantine. Numerical simulations of the mean-field model that does not include age-specific differences in disease parameters and contacts has shown that for the same mean incubation and recovery periods, increasing the number of stages of incubation period makes the epidemic reach its peak and die down faster, while increasing the number of stages in the recovery period leads to a significant increase in the maximum total numbers of infected and critical care cases.

Age structure has a major effects on how effective the quarantine is in “flattening the curve”, i.e. in reducing the number of hospitalised and critical care cases. In terms of optimal quarantine timing, if it is introduced for only 8 weeks, introducing it sooner will have a smaller effect on reducing the second peak of infection. In fact, it may even be possible that the second peak will be smaller than the first, but we only observed this for quarantines that were introduced later in the epidemic. In terms of critical care cases, there is also a notable variation between different regions in terms of whether the second peak is smaller than the first, and by how much it is reduced compared to the case with no quarantine, depending on when a fixed-duration quarantine is introduced. Effectively, this suggests that if the quarantine is introduced for some fixed duration, the timing of its introduction should be adjusted for each particular region to achieve maximum effect, or it should be kept for a sufficiently long time. We have also investigated the potential impact of quarantine duration when it is introduced simultaneously in different regions 8 weeks after the start of an epidemic. The results suggest that changing the quarantine duration from 8 to 12 or 16 weeks does shift the timing of the second peak, but has very little effect on the magnitude of this second peak in terms of either the total number of infected, the number of critical care cases, or deaths. The biggest percentage reduction in these numbers was observed for Brighton and Hove, the region with the youngest population among the three we considered.

The model presented in this paper can be made more realistic by including some additional features of disease dynamics, and further details of its progression. This includes relaxing the assumption about equality of latency (time from infection to becoming infectious) and incubation (time from infection to displaying symptoms) periods and considering them as two independent parameters; inclusion of additional compartments, such as deaths outside hospitals; analysis of some specific settings in which disease is transmitted, such as care homes, with their particular age-specific contacts. Similarly to other SIR- and SEIR-type models, our model has an underlying assumption that disease confers a life-long immunity. At this point, the data on potential secondary COVID-19 infections are rather limited, but should it become established that immunity from COVID-19 is temporary, this can be easily incorporated into a model by including direct transitions from the recovered classes back to susceptibles. Model parameters can also be further refined once more detailed age-stratified data on cases, deaths, and progression of patients in hospitals become available, though we expect the main qualitative conclusions of our analysis to hold.

## Data Availability

All data are contained within the manuscript.

